# Association of Gout with Brain Reserve and Vulnerability to Neurodegenerative Disease

**DOI:** 10.1101/2022.11.09.22282119

**Authors:** Anya Topiwala, Kulveer Mankia, Steven Bell, Alastair Webb, Klaus P Ebmeier, Isobel Howard, Chaoyue Wang, Fidel Alfaro-Almagro, Karla Miller, Stephen Burgess, Stephen Smith, Thomas E Nichols

## Abstract

**Objectiv:** To assess the associations between gout, brain structure, and neurodegenerative disease incidence.

**Methods:** Using observational and Mendelian randomization analyses we investigated causal relationships between gout and brain health. Exposures included gout diagnosis (from self-report, linked health records and death records) and genetically proxied gout and serum urate. Outcomes were neuroimaging markers of brain structure and neurodegenerative disease incidence (ascertained through self-report, health records and death reports). Cox proportional hazards models were used to examine time to neurodegenerative disease diagnosis.

**Results:** 11,735 UK Biobank participants (mean age 55.5±8.0 years and 50.5% female) had a diagnosis of gout (n=1165 in MRI subset). Dementia was ascertained in 3126 individuals over a mean follow-up time of 12.4±1.9 years. Gout patients had smaller global and regional brain volumes and higher brain iron markers. Genetic associations mirrored observational associations. Genetically proxied gout associated with lower global grey matter volume (beta= -0.05[-0.08 to -0.01]). Participants with gout had higher incidence of all-cause dementia (hazard ratio (HR)=1.60, 95% confidence interval (CI) [1.38-1.85]), Parkinson’s disease (HR=1.43[1.15 to 1.79]), and probable essential tremor (HR=6.75[5.69 to 8.00]). Risks were strongly time dependent, whereby associations with incident dementia were highest in the first 3 years after gout diagnosis (HR=7.40[4.95 to 11.07]).

**Conclusions:** These findings suggest gout is causally related to several measures of brain structure. Lower brain reserve amongst gout patients may explain their higher vulnerability to multiple neurodegenerative diseases.

**Key points:** *What is already known on this topic?:*

- Studies of neurodegenerative disease risk in gout are contradictory.
- Relationships with neuroimaging markers of brain structure, which may offer insights, are uncertain. What this study adds?

- In this prospective cohort study gout was associated with smaller brain volumes and higher incidence of multiple neurodegenerative diseases.
- Mendelian randomization analyses suggested gout is causally related to brain structure.

How might this study affect research, practice or policy?

- Our findings emphasise the importance for clinicians of assessing for motor and cognitive impairments amongst gout patients, particularly in early years after diagnosis.

## Introduction

Gout is the most common inflammatory arthritis affecting approximately 1-4% of the population [1]. The clinical syndrome is characterised by acute flares of joint pain and swelling, resulting from deposition of monosodium urate crystals in joints and peri-articular tissues, initiating an acute inflammatory cascade. In contrast to multiple other organ systems [2], classically the brain is not thought to be affected. However emerging studies have cited contradictory associations between hyperuricaemia and neurodegenerative diseases [3,4]. Some describe a lower risk of dementia, particularly Alzheimer’s disease, in hyperuricaemia [3-5]. Antioxidant properties of uric acid have been proposed as a potential mechanism for this neuroprotection [6]. At the same time, hyperuricaemia and gout have been linked to higher stroke risk [7]. Clarifying the impact on the brain is vital given that hyperuricaemia is a treatable target.

Insights into relationships between gout and neurodegenerative disease could result from examining links with brain structure, as yet unexplored. MRI markers provide quantitative, sensitive intermediate endophenotypes for neuropsychiatric disease [8]. A few studies have investigated associations between serum urate and a handful of biomarkers for stroke and dementia. Most have found no association [9,10]. However, to date no studies have examined gout. Urate analyses have been small (n<2500), not accounted for many potential confounding variables, and explored only a few aspects of brain structure, while using solely observational approaches.

We performed the first investigation of neuroimaging markers in patients with gout. Observational and Mendelian randomization (MR) approaches were combined for stronger causal inference. Furthermore, we explored relationships between gout and relevant neurodegenerative diseases. The purpose of this study was to assess whether associations between gout and brain structure would provide insights into relationships with neurodegenerative disease risk.

## Methods

### Study population

Participants were from UK Biobank (UKB) study [11], which recruited volunteers aged 40-69 years in 2006-10. A subset underwent imaging which included brain MRI (∼50,000 scanned to date). Participants underwent imaging at three centres (Newcastle, Stockport or Reading) with identical Siemens Skyra 3T scanners (software VD13) using a standard 32-channel head coil. Details of image pre-processing and quality control pipelines are described elsewhere [12]. Participants with complete data were included (**SFig 1**).

### Exposure measurements

Gout diagnoses were algorithmically defined from UKB’s baseline assessment data collection, linked data from Hospital Episode Statistics, primary care and death records (https://biobank.ndph.ox.ac.uk/showcase/refer.cgi?id=460). Individuals who developed gout after dementia were excluded to minimise reverse causation. Serum urate (in μmol/L) was measured using a Beckman Coulter AU5800 at study baseline in all participants.

### Outcome measurements

#### Brain markers

We used 2138 summary image-derived phenotypes (IDPs) representing distinct measures of brain structure from the following modalities: T1-weighted and T2-weighted-FLAIR structural imaging, susceptibility-weighted MRI, diffusion MRI. These are outputs of a dedicated processing pipeline and available from UKB [13]. IDPs included: whole brain and cerebrospinal fluid (CSF) volumes, cortical volumes, surface area, thickness, T2-FLAIR white matter hyperintensities (WMH) volumes [14], brain iron deposition metrics (T2* and magnetic susceptibility [15]), white matter microstructural measures (**SMethods**).

The precise spatial distribution of associations between gout and grey matter volume (T1-weighted images) across the brain was investigated using FSL-VBM [16] (**SMethods**), a method to compare grey matter volume in each 3D volume element (voxel) of the structural image, after adjusting between individuals for estimated total intracranial volume.

#### Neurodegenerative disease

Cases were algorithmically defined from UKB’s baseline assessment data collection (including self-report), linked data from Hospital Episode Statistics, primary care and death records (https://biobank.ndph.ox.ac.uk/showcase/refer.cgi?id=460). Incident cases (diagnosed any time after baseline) were included. Prevalent cases (diagnosis before baseline) and those who developed neurodegenerative disease prior to gout were excluded, to lessen reverse causation. The primary outcome was all-cause dementia. In planned subgroup analyses, Alzheimer’s and vascular dementia were examined.

### Covariates

Selected *a priori* confound covariates were identified on the basis of literature supporting their impact on both gout and MRI markers: all image-related confounds, age, age^2^, sex, educational qualifications, Townsend Deprivation Index, household income, historical job code, waist-hip-ratio, alcohol, smoking, and diuretic use (*Model 1*). Additional adjustment for potential consequences of hyperuricaemia (potentially on the causal pathway) included: blood pressure, cholesterol, cystatin C, creatinine, diabetes mellitus, urate-lowering therapy (ULT), and chronic kidney disease (*Model 2)*. **SMethods** describes measurement details.

### Genetic variants

Mendelian randomization was performed for two related exposures: 1) serum urate (two-sample), and 2) gout (one-sample). Genetic instruments for urate (n=179) were selected based on the sentinel variants at genome-wide significant loci (p<5×10^−8^) reported in the largest publicly available European ancestry genome-wide association study [17] (**STable 1**). Genetic instruments for gout (n=12) were selected from the largest GWAS of gout versus asymptomatic hyperuricaemia (aiming to distinguish high urate from inflammatory components of gout) [18]. Genetic associations with brain phenotypes were obtained from the largest GWAS of brain imaging phenotypes [19].

### Statistical analysis

All analyses were performed in R (version 4.1.2) unless otherwise stated.

#### Observational analyses

Multivariable linear regression models assessed the relationship between gout/urate and each of 2138 IDPs as outcomes. Brain measures and urate were quantile normalized resulting in Gaussian distributions with mean zero and standard deviation one. To account for multiple testing, Bonferroni and (separately) 5% false discovery rate (FDR) multiple comparison corrections were applied (2138 tests). Effect sizes were compared with those of age (**SMethods**). For VBM, the Big Linear Model toolbox (**SMethods**) was used to perform mass univariate ordinary least squares regression (parametric inference) voxelwise.

Cox proportional hazards models were used to estimate associations between gout/urate and neurodegenerative disease incidence. Length of follow-up was calculated as the interval between the origin and either date of event (first disease code) or censoring (date of death or last date of data collection – 2^nd^ January 2022). Origin was the baseline date, except for participants who developed gout after baseline, where gout diagnosis date was used. The impact of time to gout diagnosis on neurodegenerative disease incidence was explored by including the interval between study baseline and gout diagnosis date as a fixed covariate. Relationships between the urate level and dementia were assessed using linear (fixed effects) and non-linear regression models. For the latter urate was categorized into quintiles and restricted cubic splines (5 knots) applied [20]. Non-linearity was formally tested (H0: β2=β3=…=βk−1= 0) with an F-test. Assumptions were checked as described in **SMethods**. Associations between gout and neurodegenerative disease incidence accounting for the competing risk of death were assessed using the subdistribution method [21]. In sensitivity analyses participants with asymptomatic hyperuricaemia (defined by serum urate levels >357 μmol/L in females and >428 μmol/L in males [22]) were excluded from the control group in gout analyses.

#### Genetic analyses

We used MR to investigate whether causal relationships could underpin the observational associations we found with brain structure (**SMethods** for further details including power calculation). One-(gout) and two-sample (urate) linear MR analyses using summary statistics from European participants were conducted using R packages *MendelianRandomization* (version 0.5.1) and *TwoSampleMR* (version 0.5.6). Variant harmonization ensured that association between SNPs and exposures, and between SNPs and outcomes, reflected the same allele. No proxies were used given the availability of the vast majority of SNPs across datasets. Several robust MR methods were used to evaluate the consistency of the causal inference. To adjust for multiple testing, Bonferroni and false discovery rate (FDR, 5%) corrected p values were calculated across 20 tests.

#### Post hoc analyses

To investigate whether positive associations between urate and a few IDPs could be the result of residual confounding, models were stratified by income. After finding associations between gout and cerebellum, midbrain and striatum, additional relevant clinical outcomes were also examined *post-hoc*: incidence of Parkinson’s disease and incidence of other extrapyramidal movement disorders (ICD10 G25). The latter is likely dominated by essential tremor given its prevalence, but also includes drug-induced tremor, myoclonus, akathisia. We refer to this diagnosis henceforth as probable essential tremor.

## Results

### Participant characteristics

11,735 participants (1165 within the imaging subset) had a diagnosis of gout. Medical professionals made the majority of diagnoses (31.1% primary care, 18.5% hospital admission). A minority (15.2%) were solely self-reported. 30.8% of gout patients reported current use of ULT at assessment. Gout sufferers were older and comprised a greater proportion of males. Relationships between baseline urate and demographic variables differed by sex (**STable 2**). In males, urate was positively correlated with alcohol intake and lower socioeconomic status. This was not the case in females. Mean baseline serum urate amongst asymptomatic individuals was 305.0±76.7 μmol/L. During follow up 3126 participants developed dementia and 16,422 individuals died. Deaths amongst gout patients were more than double those of controls (11% vs. 5%).

### Observational analyses

#### Brain markers

Gout and higher urate were associated with multiple measures of brain structure (**Fig 1 & STable 3**). Associations were generally robust after additional adjustment for urate-lowering therapy and consequences of hyperuricaemia. Urate inversely associated with global brain volume, and also grey and (separately) white matter volumes, with corresponding higher cerebrospinal fluid volumes. To contextualize the effect size, we related the differences to the cross-sectional effects of age (discounting nonlinear effects of age). The effect of gout on global grey matter was equivalent to 2-year greater age, assuming all other potentially confounding factors were held constant.

**Figure 1:**
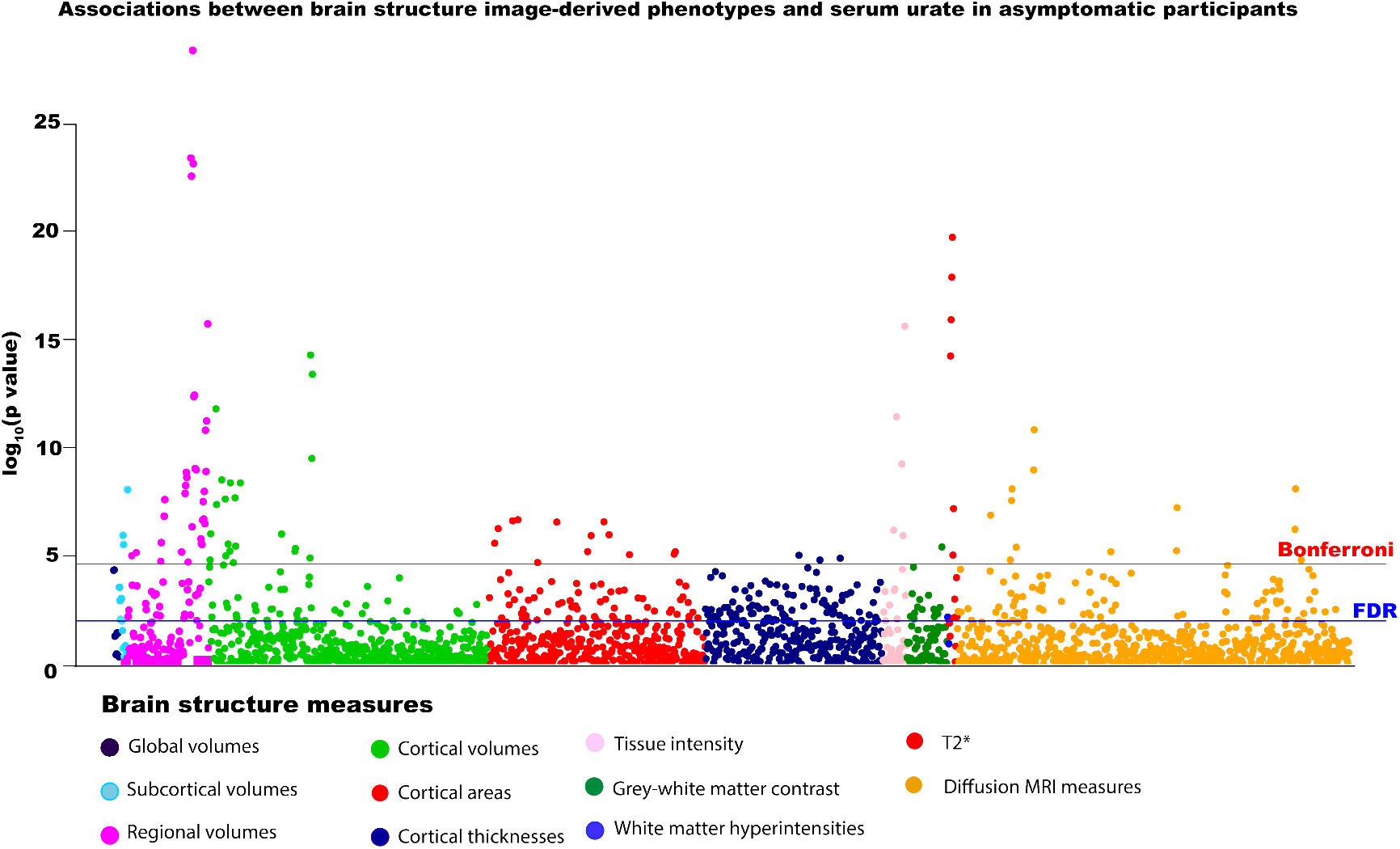
Associations between serum urate and brain structure measures in asymptomatic participants (n=33,367). Models adjusted for: all image-related confounds, age, age^2^, sex, educational qualifications, Townsend Deprivation Index, household income, historical job code, waist-hip-ratio, alcohol intake, smoking status, diuretic use. Blue line represents False Discovery Rate (FDR) threshold (5%) = 9.58×10^−3^, red line represents Bonferroni threshold on 2138 tests = 2.34×10^−5^).

There were highly significant differences in regional grey matter volumes, particularly of mid- and hindbrain structures, such as cerebellum, pons and midbrain in gout and high urate (**Fig 2**). Cerebellar differences were focused on the posterior lobe, particularly inferiorly. Subcortical differences centred on the nucleus accumbens, putamen and caudate. Significant differences were also observed in white matter tract microstructure in the fornix, the major output of the hippocampus. These included lower fractional anisotropy and higher mean diffusivity. Notably, there were no significant associations with hippocampal volume or WMH volumes. Gout and higher urate significantly associated with markers of higher iron deposition (lower T2* and higher magnetic susceptibility) of several basal ganglia structures, including bilateral putamen and caudate. Higher urate (but not gout) was positively associated with cuneal and frontal gyrus volumes in model 1, but not in model 2 or in stratified analyses amongst solely highest earners (**STable 4**).

**Figure 2:**
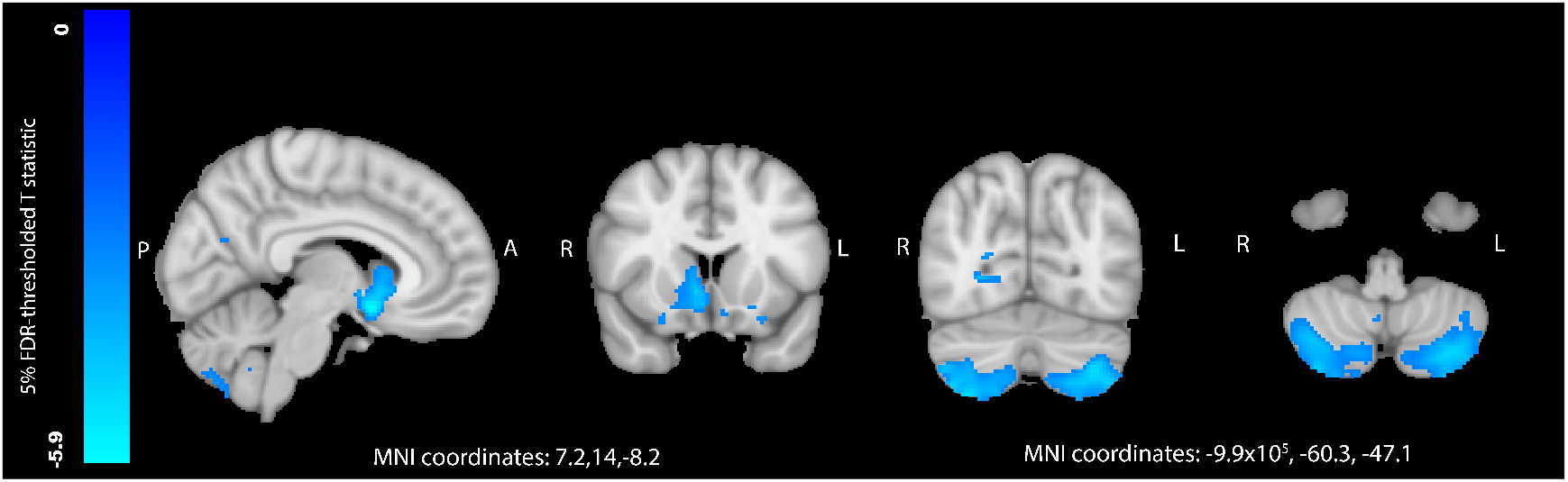
Differences in regional grey matter volume between participants with gout (n=1165) and controls (n=32,202), as analysed by voxel-based morphometry. Blue regions represent areas where participants with gout had significantly less grey matter. T statistics are thresholded at a 5% False Discovery Rate (0.0013 threshold on uncorrected p values). Models adjusted for: age, age^2^, alcohol units weekly, imaging site, smoking status, waist-hip-ratio, total household income. Abbreviations: FDR – false discovery rate, L – left, R – right, A – anterior, P – posterior, MNI – Montreal Neuroimaging Institute.

#### Neurodegenerative disease

Gout associated with a higher incidence of dementia (average over study HR=1.60[1.38 to 1.85]). The risk was time varying, highest in the first 3 years after gout diagnosis (HR=7.40[4.95 to 11.07]) and then decreasing (**Fig 3 & STable 5**). We explored possible reasons for this temporal gradient. Accounting for the competing risk lowered hazard ratios but did not alter the pattern of associations (first 3 years HR=3.53[2.31 to 5.38])(**STable 6**). Excluding controls with asymptomatic hyperuricaemia (average HR=1.59[1.42 to 1.78]) or additionally controlling for possible consequences of hyperuricaemia (average HR=1.54[1.27 to 1.87]) made little impact. Receiving a gout diagnosis later in the study (independently from age) was associated with a slightly higher dementia incidence (average HR=1.03[1.01 to 1.05]). ULT at baseline assessment was not linked to dementia incidence (HR -0.25[0.59 to 1.04]).

**Figure 3:**
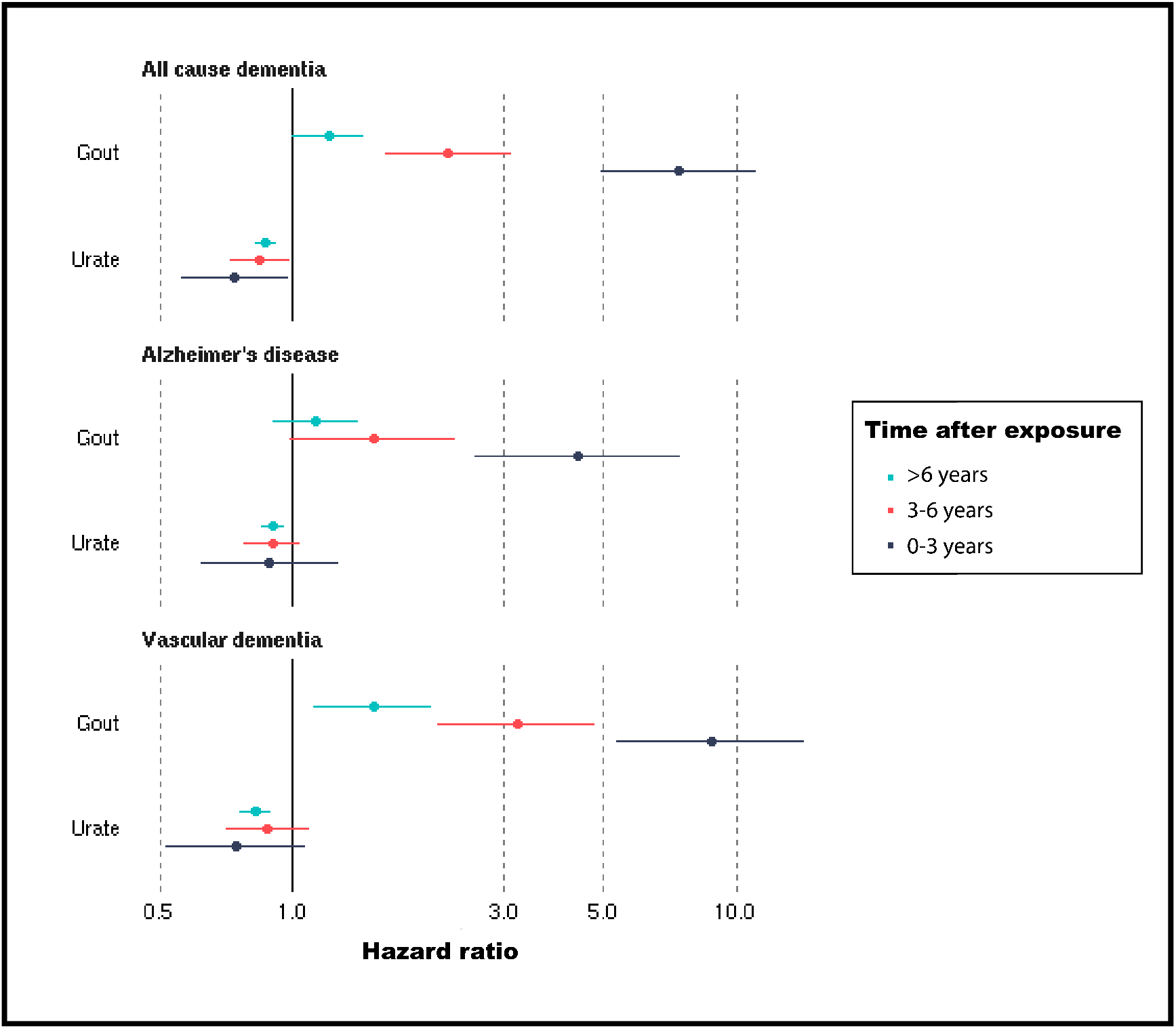
Hazard ratios of incident dementia for gout (N=303,149, 3126 dementia cases) and serum urate in asymptomatic participants (N=247,328, 2454 dementia cases). Estimates represent hazard ratios over different time periods after gout diagnosis or urate measurement, generated from Cox proportional hazards models, adjusted for: age, age^2^, sex, Townsend Deprivation Index, educational qualifications, household income, historical job code, smoking, alcohol intake, waist-hip-ratio, diuretic use.

Risks were higher for vascular dementia (average HR=2.41[1.93 to 3.02]) compared to all-cause dementia, but not for Alzheimer’s disease (average HR=1.62[1.30 to 2.02]). Again, there was a strong time-dependence to the risks, particularly for vascular dementia. Amongst asymptomatic individuals, in the linear model there was an inverse association between urate and dementia incidence (HR=0.85, 95% CI: 0.80 to 0.89), with no time dependence. Using restricted cubic splines a non-linear relationship between urate and dementia incidence was observed (p for non-linearity <0.0001). Both low and high urate levels associated with a higher dementia incidence (**SFig 2**).

Given the associations with cerebellum and striatum measures, we examined relevant phenotypes *post-hoc*. Gout was associated with higher incidence of both Parkinson’s disease (HR=1.43[1.15 to 1.79]) and probable essential tremor (HR=6.75[5.59 to 8.00]). Amongst asymptomatic individuals, there was a linear inverse relationship between urate and incident Parkinson’s disease (HR=0.89[0.84 to 0.95], p for non-linearity=0.3), but not with probable essential tremor (HR=0.95[0.89 to 1.02]).

### Genetic analyses

Genetic associations mostly mirrored observational ones. Both genetically predicted gout and serum urate significantly associated with regional grey matter volumes, including cerebellar, midbrain, pons and brainstem. There were also significant associations with higher magnetic susceptibility in the putamen and caudate, markers of higher iron (**Fig 4**). However, whilst genetically predicted gout significantly associated with global grey matter volume, urate did not. All associations survived FDR correction for multiple testing. Pons, brain stem and the magnetic susceptibility associations additionally survived Bonferroni correction. Robust MR methods gave broadly consistent estimates, albeit with wider CIs (**STable 7**).

**Figure 4:**
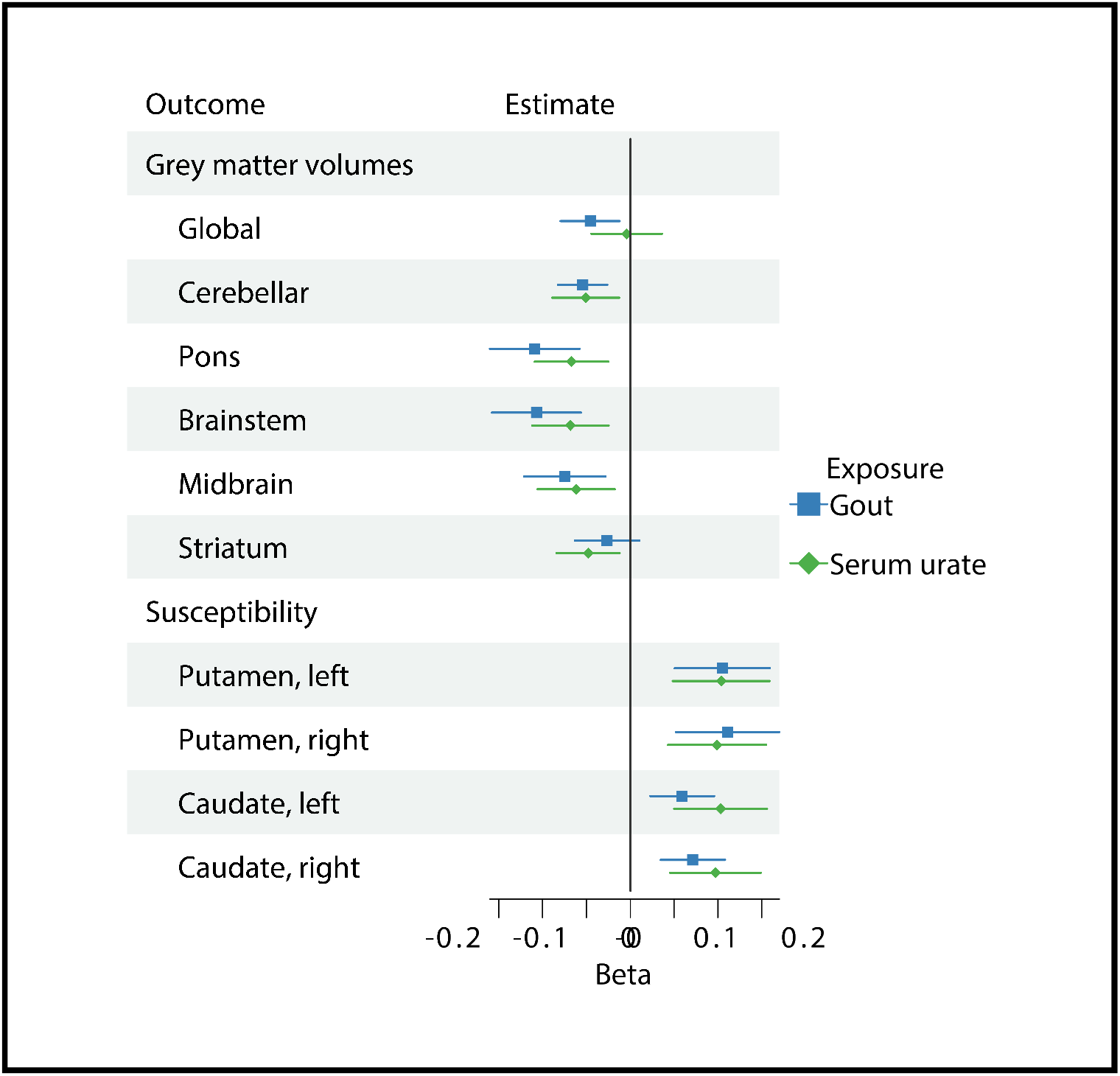
Mendelian randomization estimates for the association of genetically predicted gout and serum urate and brain imaging phenotypes. Genetically-predicted gout was instrumented using 12 SNPs (one-sample MR). Genetically-predicted serum urate was instrumented using 179 SNPs (two-sample MR). Beta estimates represent inverse-variance weighted estimates for a 1mg dl^−1^ increase in serum urate, or gout versus asymptomatic hyperuricaemia. Magnetic susceptibility measures are derived from quantitative susceptibility maps, with higher values indicating higher iron. Abbreviations: IDP – image-derived phenotype, SNPs – single nucleotide polymorphism, LCI – lower confidence interval, UCI – upper confidence interval.

## Discussion

In this UK prospective cohort study, participants with a history of gout had smaller global and regional brain volumes and higher brain iron. MR analyses suggested gout was causally related to brain structure. Gout was associated with a higher incidence of several neurodegenerative diseases, particularly in the first three years after diagnosis. Lower brain reserve in gout patients may explain their vulnerability to dementia.

To our knowledge, there are no previous neuroimaging studies of gout. Investigations examining urate are small and, with two exceptions, examined solely markers of cerebrovascular disease [23]. One study reported higher white matter atrophy with hyperuricaemia [24], and another a null association with hippocampal volume [9]. We replicated these findings, but did not duplicate reported positive relationships between urate and WMH [25]. Methodological differences may be responsible. In UKB, WMH volumes were calculated automatically whereas previous studies used visual ratings. We found novel associations with global grey matter and cerebellar (motor and non-motor regions [26]), brainstem and striatal brain volumes. Gout patients had higher incidence of probable essential tremor, previously linked to cerebellar atrophy [27,28]. Associations with markers of higher iron in basal ganglia with hyperuricaemia mirror findings in aging, Alzheimer’s and Parkinson’s diseases [29,30], and recently alcohol consumption [31]. Our MR findings provide support for causal relationships underlying the associations.

The mechanism(s) underlying how gout affects brain volume is unclear. Hyperuricaemia has been linked to arterial stiffness [32], and associated with brain microvascular damage [33], which may improve with allopurinol treatment [34]. Alternatively, toxic metabolic pathways may be responsible [35,36]. It is unclear whether, at least in healthy individuals, urate can cross the blood brain barrier [37]. High levels of iron in basal ganglia could result from inflammatory processes in gout [38,39], or conversely, higher ferritin (a blood measure of iron) secondary to diet, could lead to higher urate levels [38].

In this sample gout associated with higher incidence of dementia, a finding at odds with previous claims of protective effects [23,40]. A recent small meta-analysis found no evidence for an association with gout overall, but cited a possible protective effect on Alzheimer’s disease based on two studies [5]. Whilst gout associated with smaller global brain volumes, we did not observe associations with classical imaging markers of Alzheimer’s disease [41] or vascular dementia [42] such as hippocampal volume [43] or white matter hyperintensities. Instead as gout played a causative role in multiple neurodegenerative pathologies, we propose a brain vulnerability model (**Fig 5**). A similar phenomenon is well recognized with delirium [44]. Global brain volumes are commonly conceived as a proxy for brain reserve, structural characteristics of the brain that allow some people to better cope with brain pathology before cognitive changes emerge [45]. The strong temporal gradient in risk of neurodegenerative disease after gout is interesting. We propose three hypotheses. First, death is a competing risk. Second, there may be a selective detection bias. Receiving a gout diagnosis may result in more frequent medical review initially, when cognitive problems could be noticed. Third, the inflammatory response in a gout flare may induce a stepwise global decline that may precipitate acute cognitive decline.

**Figure 5:**
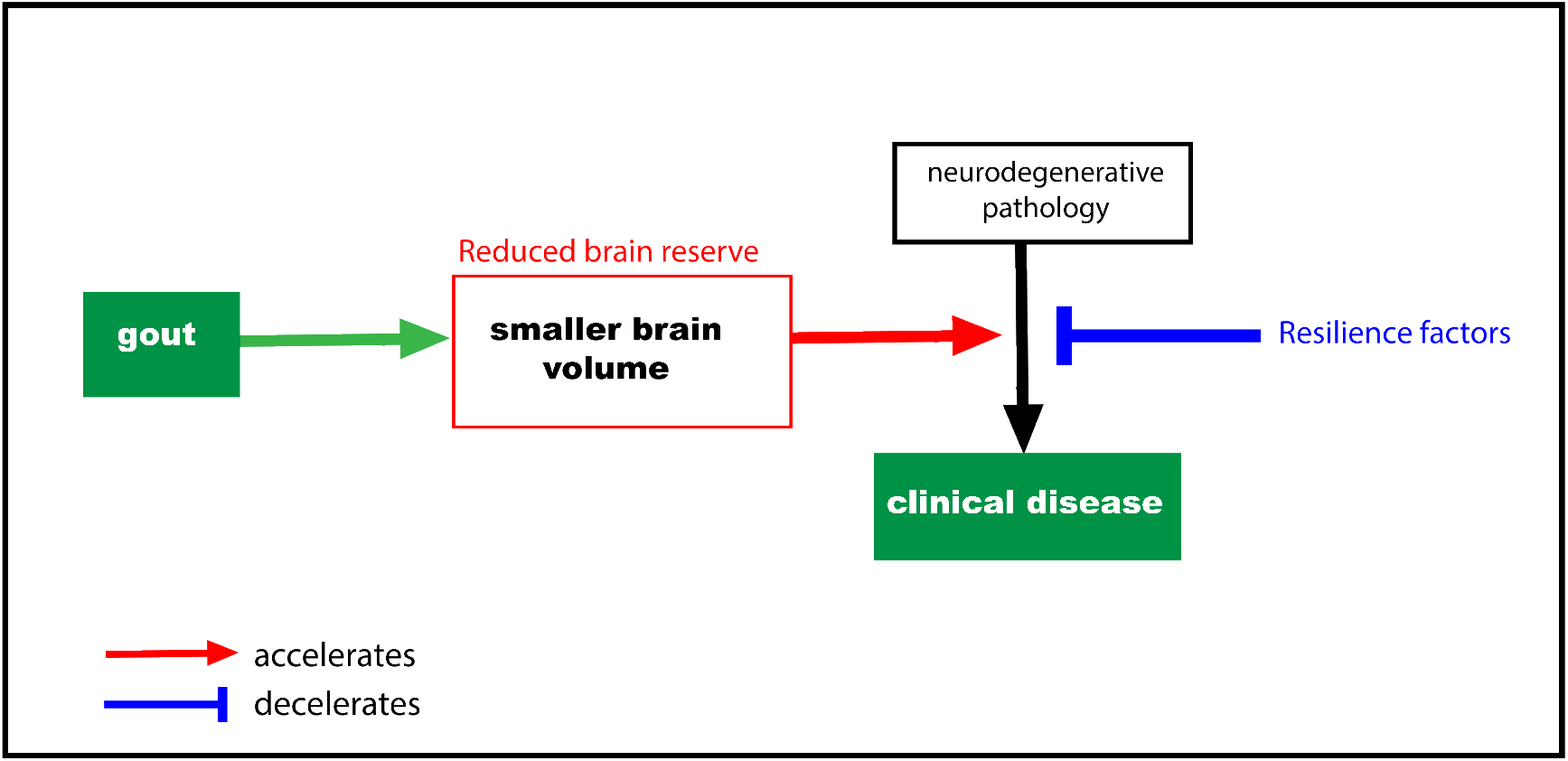
Hypothesised model for the impact of gout on neurodegenerative disease. Gout is causally related to smaller brain volume, a proxy for cognitive reserve. Reduced cognitive reserve indicates a reduced ability for individuals to tolerate neurodegenerative pathology before symptoms ensue.

The relationship between baseline urate and dementia incidence was non-linear. As others have previously raised, associations with a single urate measurement, which may not reflect long-term exposure [46], should be interpreted cautiously. Reduced risks of dementia with hyperuricaemia have been previously reported [3], including in an earlier analysis of UKB [4]. Given brain associations with urate and gout were consistent, we hypothesise collider bias may be causing a spurious protective association for urate [47]. Both higher urate (in affluent individuals), and absence of dementia increase likelihood of UKB recruitment. A recent MR study found higher urate linked with increased Alzheimer’s supports this hypothesis [48].

These findings hold potential implications for gout patients and also for clinicians. Prophylaxis for gout flares may be justified on the basis of brain health in addition to joint and cardiovascular health, although interventional trials will be needed to test this. Patients with gout should be monitored for cognitive and motor symptoms of neurodegenerative disease given their increased risk, especially in the early period after diagnosis.

### Strengths and limitations

The large size and statistical power enabled triangulation of observational and MR approaches for stronger causal inference. A comprehensive range of brain metrics were examined, whilst adjusting for multiple confounds and multiple testing. Other key strengths include the prospective cohort, examination of relevant clinical phenotypes in the same sample and time to event analyses accounting for competing risks.

This study has several limitations. First, UKB is a self-selective cohort [49]. Second, measurement of serum urate was cross-sectional and may not reflect chronic exposure [50]. Third, MRI data was at a single time point so we cannot infer changes in brain structure. Fourth, case identification may be subject to ascertainment bias, likely skewed to more severe cases, which could bias associations towards the null. Fifth, there may be diagnostic confounding between Parkinson’s disease and essential tremor. Sixth, gout covaries with obesity and alcohol. However, we controlled for these factors, performed MR analyses, and we have previously reported brain structure associations with alcohol in a markedly different spatial distribution in the same sample [51]. Seventh, MR relies on a set of unverifiable assumptions, which we tried to assess where possible. Eighth, sample overlap between gout and brain imaging GWAS may bias genetic associations towards observational associations. Finally, this was not a bespoke study with optimal measures for testing cerebellar function, for example measured gait speed or eye movements.

## Conclusions

In this population-based sample, participants with gout had smaller global and regional grey matter volumes. Lower brain reserve may explain vulnerability of gout patients to neurodegenerative disease. Clinicians should be vigilant for motor and cognitive problems in gout patients.

## Supporting information

SMethods

SFigures

STable 1

STable 2

Stable 7

STable 6

STable 5

STable 3

STable 4

## Data Availability

All data in the present study are available upon successful UKB application or publically available summary statistics

## Acknowledgements

We wish to thank Maria Christodoulou and the University of Oxford Statistics Consulting service for providing advice on the survival model analyses.

## Data sharing

All data is available upon successful application by bona fide researchers through UK Biobank.

