## Supplementary material for "Association of Gout with Brain Reserve and Vulnerability to Neurodegenerative Disease": SMethods

**MRI acquisition**

As part of the protocol [1] T1-weighted images were obtained using an MPRAGE sequence: TR=2000ms, TE=2.0ms, 208 sagittal slices, flip angle=8°, FOV=256mm, matrix=256x256, slice thickness=1.0mm (voxel size 1x1x1mm). Diffusion images were obtained using a spin-echo echo-planar sequence with 10 T2-weighted baseline volumes, 50b = 1000 s mm^-2^ and 50 b=2000 s mm^-2^ diffusion weighted volumes, with 100 diffusion-encoding directions and 2mm isotropic voxels. Resting state functional MRI was acquired with the following parameters: TE=39ms, TR=735ms, MB=8, R=1, flip angle=52°, 490 time points, 2.4x2.4x2.4mm voxel size.

**Covariate measurement**

The full set of image-related confounds (>600) included intracranial size, head motion, scanner table position, imaging centre and date [2]. QR decomposition was used to remove linear dependence between variables. Smoking status was reported as categorical variable: never/previous/current. Waist-hip ratio and blood pressure were assessed at baseline. Weekly alcohol consumption (in UK units) was estimated at baseline by summing across beverage types as previously described [3]. Educational qualifications and total household income were self-reported in categories. Townsend Deprivation Index was used as a continuous measure of deprivation based on census information. Historical job type was coded according to the Standard Occupational Classification 2000 [4]. Major groups with sufficient numbers in UKB were dummy coded (managers and senior officials, professional occupations, associate professional/technical occupations, administrative/secretarial occupations). Urate-lowering therapy or gout treatment (including allopurinol, colchicine, sulphinpyrazone) and diuretic medication (yes/no) were ascertained at baseline and imaging visit by verbal interview by a nurse. Chronic kidney disease (presence of ICD code N18) was coded as a binary variable. Diabetes was coded as a binary variable (presence of an ICD code for non-insulin dependent, insulin dependent, or non-specified diabetes, E10, 11&14). Total and high-density cholesterol, creatinine and cystatin C (markers of renal function) were derived from a blood sample at baseline.

**Neuroimaging measures**

***Iron markers:*** Brain iron content was ascertained using quantitative susceptibility mapping (QSM) and T2*, both derived from susceptibility-weighted magnetic resonance imaging (swMRI) data. T2* reflects differences in tissue microstructure related to iron (sequestered to ferritin) and myelin, and correlates with post-mortem estimates of iron deposits in brain grey matter [5]. Susceptibility reflects the net (sequestered and non-sequestered) content of susceptibility-shifting sources like iron and myelin. Two distinct and complementary metrics of brain iron deposition were used, T2* and quantitative susceptibility mapping (QSM), to produce image-derived phenotypes (IDPs). Whilst these metrics are coupled, consistent findings across the two will provide greater evidence that iron levels are affected.

***White matter microstructure:*** Diffusion tensor imaging (DTI) measures the directional preference of water diffusion in neural tissues and allows inferences about the structural integrity of white matter tracts. Measures include:

*Fractional anisotropy* – A measure which reflects diffusion of water parallel to in relation to diffusion perpendicular to the principal fibre direction. It is widely used as a marker of white matter tract integrity.

*Mean diffusivity* – The apparent diffusion coefficient averaged over all directions.

**Voxel-based morphometry**

Relationships between alcohol use and grey matter were spatially pinpointed in a brain-wide hypothesis-free manner using FSL-VBM [6] (http://fsl.fmrib.ox.ac.uk/fsl/fslwiki/FSLVBM), an optimised voxel-based morphometry (VBM) protocol [7] carried out with FSL tools [8]. This is an objective method to compare grey matter volume (estimated total intracranial volume adjusted) between individuals in each voxel (smallest distinguishable 3D image volume) of the structural image. Only participants with usable T1 images proceeded to the VBM analysis. After brain extraction, tissue segmentation and registration, images were averaged and flipped along the x-axis to create a left-right symmetric, study-specific grey matter template. Second, all native grey matter images were non-linearly registered to this study-specific template and "modulated" to correct for local expansion (or contraction) due to the non-linear component of the spatial transformation. The modulated grey matter images were then smoothed with an isotropic Gaussian kernel with a sigma of 2 mm. We created a study specific average grey matter tissue map using unsmoothed and modulated grey matter images as per standard VBM protocol. By thresholding this map (at 0.01) a grey matter mask was created. This was used as an analysis mask.

**Statistical analyses methods**

***Big Linear Toolbox*** [9] - A missingness threshold of 80% was employed (i.e. voxels with recorded data for less than 80% of subjects were discarded from the analysis), and two T contrasts (positive and negative correlation with gout) and an F contrast were computed. A p-value threshold that controlled the False Discovery Rate (FDR) at 0.05 was generated using FSL’s FDR (<https://fsl.fmrib.ox.ac.uk/fsl/fslwiki/FDR>) and used to threshold T statistic images.

***Comparing effect sizes to age effects*** *–* Higher order age terms and age x sex interactions were orthogonalized with respect to the main linear age term. All were then entered into the regression model with total grey matter as the dependent variable, together with other covariates. The effect size for 1 unit higher urate over the study could then be compared with that for being 1 year greater in age at study baseline.

***Cox proportional hazards assumption checks -*** Influential observations were assessed by plotting deviance residuals. Proportional hazards were assessed visually using Schoenfeld residuals and formally with time interactions. Two approaches were used for variables violating the proportional hazards assumption. Time-varying coefficients were calculated for gout diagnosis by splitting the dataset into time dependent parts and then testing interactions with time [10]. For covariates not of primary interest (age, historical job, BMI and alcohol), stratified models were fitted without the constraint of non-proportionality. Separate baseline hazard functions were fitted for each stratum.

We computed the probability of being in dementia and death states (absorbing states) using the Aalen-Johansen estimate to assess whether death was a competing risk. Mean time in death state for gout cases=1.16, controls=0.40 years.

***Mendelian randomization -***

Assumptions: 1) genetic variants are robustly associated with the exposure (here gout/urate), 2) genetic variants share no common cause with the outcome (IDPs), and 3) genetic variants only affect the outcome through the exposure.

Genetic variants were selected on the basis of genome-wide significance (p<5x10^-8^) (assumption 1). Horizontal pleiotropy was assessed using robust methods (see below, assumption 3).

Robust methods

Several robust MR methods were performed to evaluate the consistency of the causal inference. Inverse variance weighted (IVW) analysis (multiplicative random effects) regresses the effect sizes of the variant-iron marker associations against the effect sizes of the variant-alcohol associations. The MR-Egger method uses a weighted regression with an unconstrained intercept to relax the assumption that all genetic variants are valid IVs (under the Instrument Strength Independent of Direct Effect (InSIDE) assumption) [11]. A non-zero intercept term can be interpreted as evidence of directional pleiotropy, where an instrument is independently associated with the outcome violating an MR assumption. The median and modal MR methods are also more resistant to pleiotropy, as they are robust when up to 50% of genetic variants or more than not, respectively, are invalid. These methods are recommended in practice for sensitivity analyses as they require different assumptions to be satisfied, and therefore if estimates from such methods are similar, then any causal claim inferred is more credible.

Multiple testing correction

To adjust for multiple testing, Bonferroni and false discovery rate (FDR, 5%) corrected p values were calculated.

Power calculations for MR analyses were based on an online calculator developed by one of the authors [12]. Based on a R^2^ of 0.077 [13] and a significance level of 0.05, the sample size of n=39,691 [14] has 79% power to detect a causal effect of 0.05 standard deviation units in imaging measure per standard deviation change in urate.

1.	Smith Stephen M A-aF, Miller Karla L. UK Biobank Brain Imaging Documentation. 2020(Version 1.8).

2.	Alfaro-Almagro F, Mccarthy P, Afyouni S, Andersson JL, Bastiani M, Miller KL, et al. Confound modelling in UK Biobank brain imaging. NeuroImage. 2021;224:117002.

3.	Topiwala A, Ebmeier KP, Maullin-Sapey T, Nichols TE. No safe level of alcohol consumption for brain health: observational cohort study of 25,378 UK Biobank participants. medRxiv. 2021.

4.	Statistics OON. Standard Occupational Classification 2000: SOC 2000. 2000.

5.	De Barros A, Arribarat G, Combis J, Chaynes P, Péran P. Matching ex vivo MRI with iron histology: pearls and pitfalls. Frontiers in neuroanatomy. 2019;13:68.

6.	Douaud G, Smith S, Jenkinson M, Behrens T, Johansen-Berg H, Vickers J, et al. Anatomically related grey and white matter abnormalities in adolescent-onset schizophrenia. Brain. 2007;130(9):2375-86.

7.	Good CD, Johnsrude I, Ashburner J, Henson RN, Friston KJ, Frackowiak RS. Cerebral asymmetry and the effects of sex and handedness on brain structure: a voxel-based morphometric analysis of 465 normal adult human brains. Neuroimage. 2001;14(3):685-700.

8.	Smith SM, Jenkinson M, Woolrich MW, Beckmann CF, Behrens TE, Johansen-Berg H, et al. Advances in functional and structural MR image analysis and implementation as FSL. Neuroimage. 2004;23:S208-S19.

9.	Maullin-Sapey TNTE. BLM toolbox for neuroimaging cluster and local usage

10.	Zhang Z, Reinikainen J, Adeleke KA, Pieterse ME, Groothuis-Oudshoorn CG. Time-varying covariates and coefficients in Cox regression models. Annals of translational medicine. 2018;6(7).

11.	Bowden J, Davey Smith G, Burgess S. Mendelian randomization with invalid instruments: effect estimation and bias detection through Egger regression. International journal of epidemiology. 2015;44(2):512-25.

12.	Burgess S. Online sample size and power calculator for Mendelian randomization.

13.	Tin A, Marten J, Halperin Kuhns VL, Li Y, Wuttke M, Kirsten H, et al. Target genes, variants, tissues and transcriptional pathways influencing human serum urate levels. Nature genetics. 2019;51(10):1459-74.

14.	Smith SM, Douaud G, Chen W, Hanayik T, Alfaro-Almagro F, Sharp K, et al. An expanded set of genome-wide association studies of brain imaging phenotypes in UK Biobank. Nature neuroscience. 2021;24(5):737-45.
