## Supplementary material for "Association of Gout with Brain Reserve and Vulnerability to Neurodegenerative Disease": SFigures

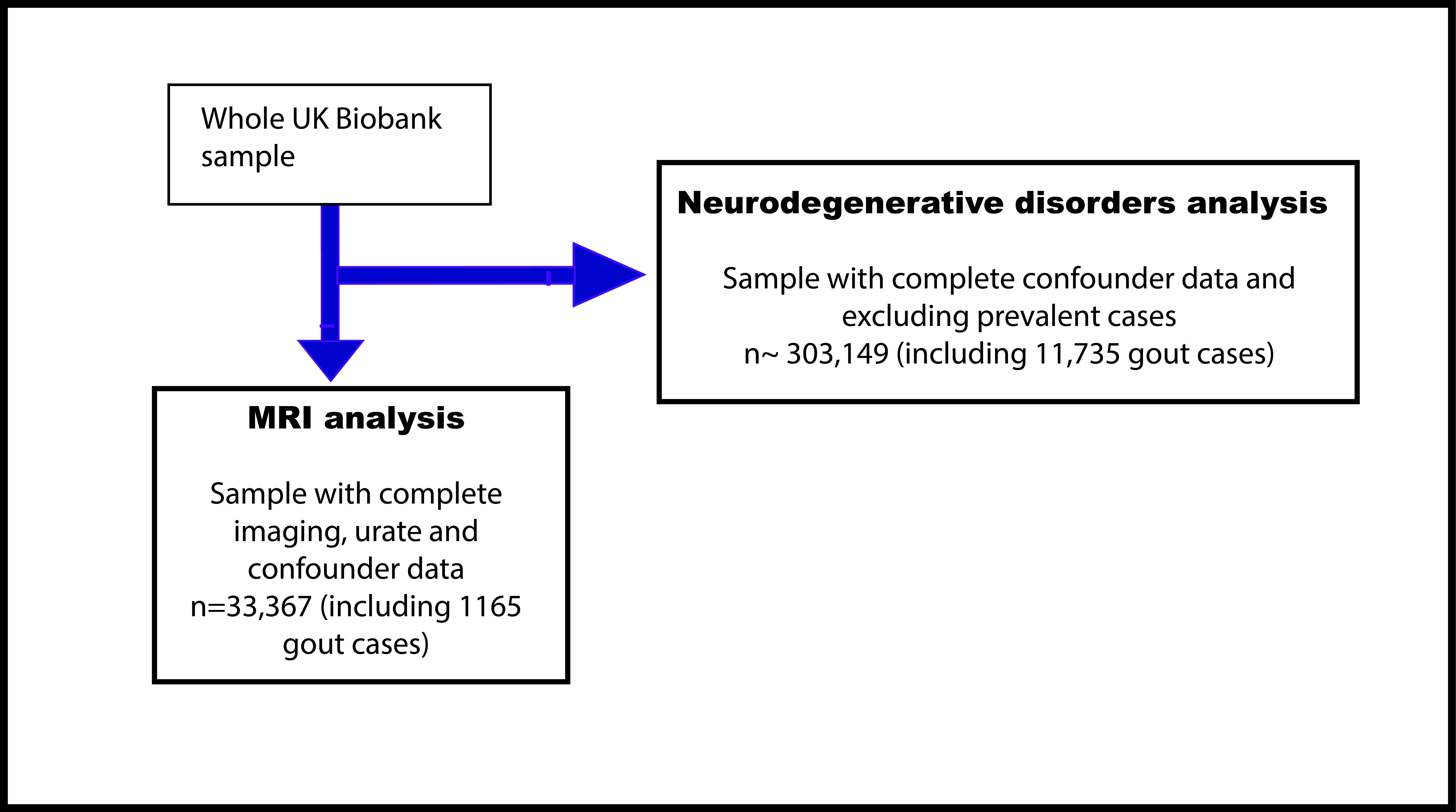

**SFigure 1: Flow chart of participants included in analyses.**

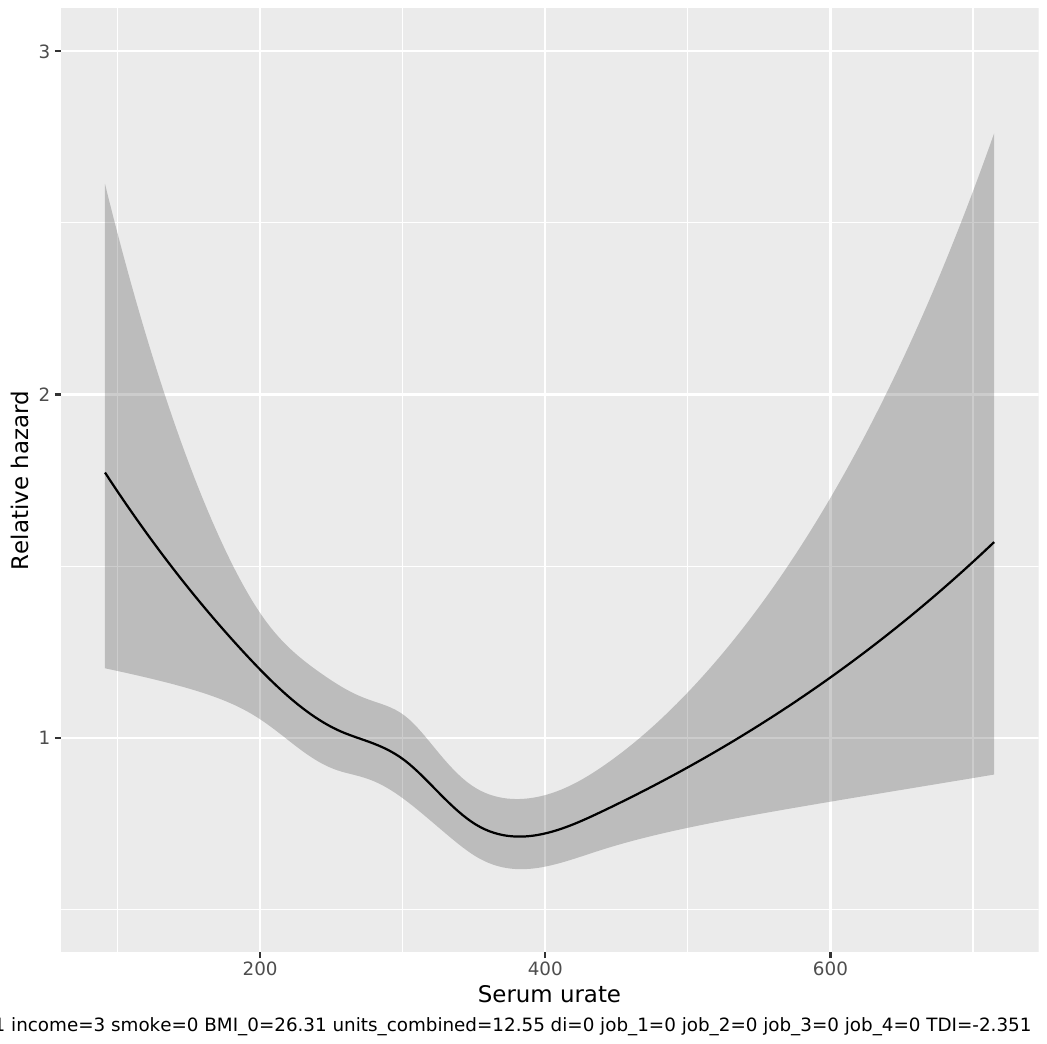

**SFigure 2: Predicted hazard of all cause of dementia according to baseline serum urate (μmol/L).** Hazards are plotted relative to that of median urate. Graphs generated from Cox proportional hazards model adjusted for: age, age^2^, sex, Townsend Deprivation Index, educational qualifications, household income, historical job code, smoking, alcohol intake, waist-hip-ratio, diuretic use. Restricted cubic splines (5 knots, quintiles) are applied to urate.
